# HIGH EMBOLIC RISK WITH ORBITAL ATHERECTOMY FOR CALCIFIED PERIPHERAL ARTERY DISEASE

**DOI:** 10.64898/2025.12.02.25341425

**Authors:** Rail Gilyazov, Barbara Batista de Oliveira, Maksim Glushkov, Paul Deegan, Jason MacTaggart, Alexey Kamenskiy

## Abstract

**Purpose:** Orbital atherectomy (OA) is used to treat heavily calcified peripheral artery disease, aiming to improve vessel compliance and reduce the need for high-pressure angioplasty and stenting. However, clinical data raise concerns regarding distal embolization, vessel injury, and uncertain long-term benefit.

**Methods:** The biomechanical and embolic effects of OA were evaluated in 10 human femoropopliteal arteries with advanced atherosclerosis and calcification. A pulsatile flow loop replicated physiologic hemodynamics. Sequential OA and balloon angioplasty (BA) were performed, followed by μCT to quantify luminal changes. Pressure and flow sensors assessed translesional gradients and flow rates. Embolic burden was captured via 100μm filters and quantified using image analysis. Arterial pulsatility was evaluated using duplex ultrasound and optical imaging.

**Results:** OA and BA produced additive luminal gains (total increase: 0.409mL) but frequently caused dissections. OA generated substantial embolic debris (mean area 0.386±1.654mm^2^), with 90% of arteries releasing fragments ≥1mm and 10% >5mm. BA added smaller debris (0.143±0.361mm^2^). Translesional pressure gradients decreased in 40% of arteries, showed no improvement in 50%, and worsened in 10%. Net flow followed a similar pattern: improved in 40%, unchanged or decreased in 60% due to dissections or distal obstruction. Inner diameter (ID) pulsatility increased from 1.5%±0.7% to 2.3%±0.5%, while outer diameter pulsatility remained unchanged.

**Conclusions:** OA improves lumen volume and ID-based pulsatility but consistently produces large emboli and vessel wall injury. These findings provide mechanistic support for the high embolic complication rates and mixed long-term outcomes observed clinically, emphasizing the importance of embolic protection and careful patient selection.

## 1. INTRODUCTION

Peripheral artery disease (PAD) affects over 200 million people globally and remains a leading cause of morbidity, limb loss, and healthcare costs in aging populations[1]. It is driven by progressive arterial narrowing from atherosclerosis, thrombosis, and calcification – most commonly in the femoropopliteal (FPA) and infrapopliteal arteries[2]. In its most advanced form, critical limb ischemia (CLI), PAD results in rest pain, tissue loss, and high amputation risk. Endovascular revascularization, typically involving balloon angioplasty (BA) and stenting, is the cornerstone of treatment. However, severe calcification impairs vessel compliance, increases the risk of recoil and dissection, and complicates device delivery and expansion[3].

Atherectomy was developed to address these challenges by mechanically debulking plaque, improving luminal gain, and enabling low-pressure BA to minimize barotrauma and reduce stent use[4]. Orbital atherectomy (OA), in particular, uses a diamond-coated, eccentrically rotating crown to sand calcified lesions while aiming to preserve vessel integrity and minimize deep injury. The device reportedly generates microparticles averaging 2.04 μm[5] – smaller than red blood cells – suggesting a reduced risk of distal embolization. OA is also thought to improve vessel compliance[6], enhancing BA effectiveness by modifying superficial and medial calcification.

Despite these theoretical advantages, real-world data have raised concerns about embolic complications and long-term efficacy. OA has been associated with higher rates of distal embolization than BA or stenting, often necessitating aspiration or surgery[7, 8]. Registry and claims-based studies have further linked atherectomy to increased amputation rates and adverse limb events compared to stenting[9], questioning its routine use[10, 11].

Yet few studies have systematically assessed OA’s biomechanical and embolic effects under controlled conditions[12]. A clearer mechanistic understanding of its impact on flow dynamics, pressure gradients, embolic load, and vessel wall behavior is needed. We hypothesized that OA yields modest luminal gain and improved compliance in calcified arteries but generates emboli large enough to obstruct distal vessels – an effect exacerbated by adjunctive BA. To test this, we used an *ex vivo* human artery flow loop with physiologic hemodynamics and μCT imaging to quantify changes in lumen volume, translesional pressure gradient, net flow, embolic burden, and pulsatility, offering mechanistic insight into clinical outcomes.

## 2. MATERIALS AND METHODS

### 2.1 Arteries

A total of 10 human FPA segments (9 males, mean age 77 ± 7 years, range 62-87 years) were obtained from tissue donors by Live On Nebraska within 24 hours of death, following next-of-kin consent for research. All donors had hypertension; 50% had diabetes, coronary artery disease, and chronic kidney disease; 70% had dyslipidemia; and 80% were former smokers while 20% were current smokers. Surrounding adipose and muscle tissues were carefully removed, and all side branches were ligated. Each artery was trimmed to a standardized length of ∼10 cm, with an effective working length of ∼8 cm to allow secure attachment to the testing rig. The rig anchored the artery at both ends, permitting imaging and hemodynamic assessments without the need for repeated mounting or repositioning, thereby minimizing handling artifacts.

Each vessel underwent three sequential assessments - at baseline, post-OA, and post-BA – to mirror standard clinical practice. OA was performed using the Diamondback 360® Peripheral Orbital Atherectomy System (Cardiovascular Systems Inc.) over a ViperWire® Advance Guidewire with ViperSlide® Lubricant, per manufacturer instructions. One pass was made at each rotational speed (60k, 90k, and 140k RPM), lasting 30-45 seconds under pulsatile flow. BA followed using semi-compliant balloons sized ∼1 mm larger than the post-OA lumen. Four balloon types were used depending on vessel diameter: Ultra-Thin™ Diamond Standard (7-8 mm, Boston Scientific), ATB Advance (6 mm, Cook Medical), and Crosstella RX (5 mm, Terumo), all inflated to nominal pressure (typically 8 atm). At each stage (baseline, post-OA, and post-OA+BA), we performed standardized assessments including μCT imaging to quantify luminal changes and detect dissections, hemodynamic measurements of translesional pressure and flow, embolic burden quantification, and evaluation of arterial pulsatility relative to baseline.

### 2.2 μCT imaging

Imaging was performed using an EasyTom S system (RX Solutions, Chavanod, France). Arteries were internally pressurized to ∼90 mmHg using shaving foam, which preserved luminal geometry and enhanced contrast during acquisition[13]. To prevent dehydration, the testing rig was wrapped in plastic film, and intraluminal pressure was continuously monitored via digital pressure gauge. Scanning parameters included 150 kV tube voltage, 100 μA current, 11 fps frame rate, and averaging of 5 frames per projection. The isotropic voxel resolution ranged from 50-55 μm. Reconstructed TIFF image stacks were processed with RX Solutions software and imported into Mimics Innovation Suite v24.0 (Materialise NV, Leuven, Belgium) for segmentation of the flow lumen, arterial wall, and calcified plaques. In addition to full-volume analysis, cross-sectional evaluations were performed slice-by-slice at predefined locations for all three stages to assess localized changes in luminal morphology, plaque removal, and dissection formation. Flow lumen cross-sectional areas were exported for each μCT slice using Mimics’ Mask Area Export function to generate longitudinal profiles. Total luminal volume was calculated from the segmented lumen mask using Mimics’ volumetric analysis tool.

### 2.3 Hemodynamic assessment

The pulsatile flow circuit was designed to replicate physiologically realistic arterial hemodynamics. At its core was the PD-1100 Pulsatile Pump (BDC Laboratories) and control module, regulated via real-time sensor feedback from the loop (Figure 1). A 2.25 L compliance chamber was integrated to simulate arterial compliance and smooth pressure waveforms. Downstream, a flow meter provided continuous volumetric flow measurements (L/min). Two 10 FR hemostasis valve Y-connectors (Rotating Male Luer Lock, Female Luer Lock Sideport) were positioned proximally and distally to the test artery, enabling device exchanges. Pressure transducers placed immediately upstream and downstream of the artery captured translesional pressure gradients under pulsatile flow. The artery was submerged in a reservoir filled with 0.9% phosphate-buffered saline (PBS), maintained at 37°C. Prior to testing, the system was calibrated using a 5 mm inner diameter PVC tube to simulate baseline hemodynamics. Flow and resistance settings were tuned to achieve physiologic FPA conditions: proximal and distal pressures of ∼120 mmHg and ∼80 mmHg, respectively, with a mean flow rate of ∼0.55 L/min under resting conditions.

**Figure 1.**
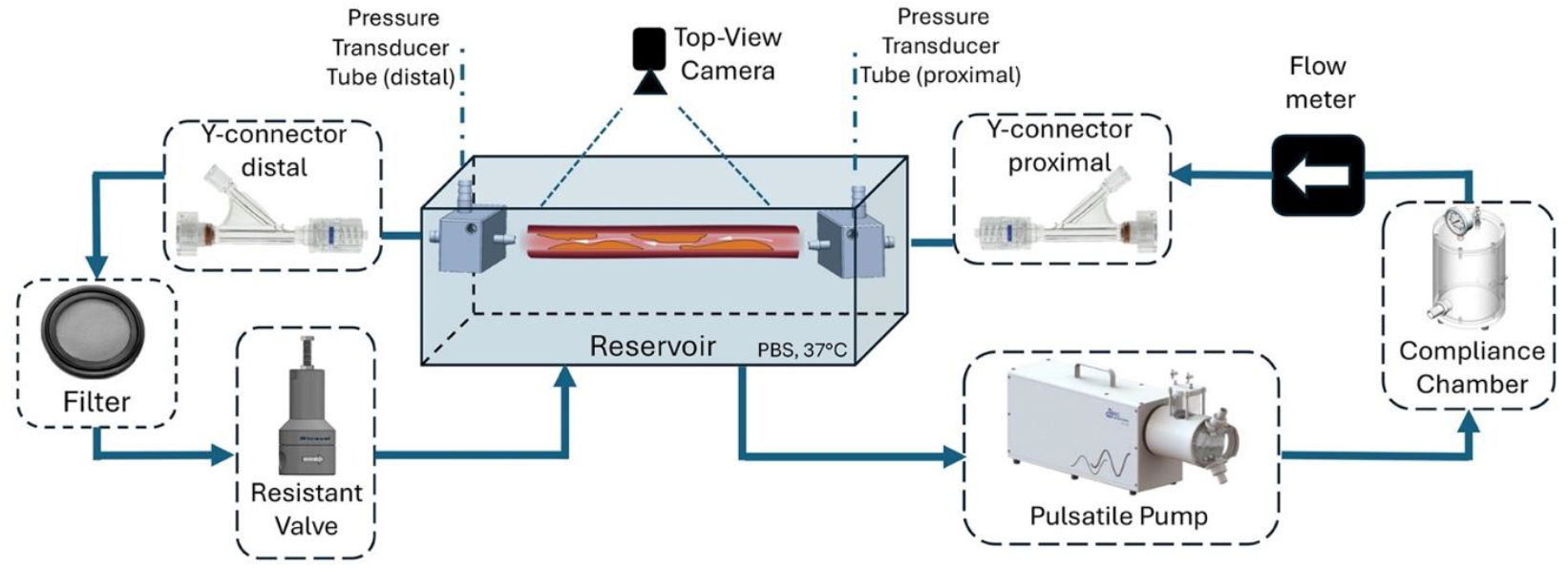
Schematic of the pulsatile flow circuit used to evaluate FPA segments. The setup includes a pulsatile pump, compliance chamber, flow meter, proximal/distal pressure transducers, top-mounted camera for diameter tracking, 100 μm embolic filter, and a distal resistance valve to simulate peripheral resistance.

To assess hemodynamic resistance, translesional mean arterial pressure gradient (ΔMAP) was computed from synchronized proximal and distal pressure waveforms acquired under continuous pulsatile flow (∼1 Hz) for 10 seconds. After waveform stabilization, instantaneous gradients (Δ*P*(*t*) = *P*_*proximal*_(*t*) − *P*_*distal*_(*t*)) were averaged over 10 stable cycles to yield ΔMAP. Volumetric flow was recorded via an ultrasonic flow meter positioned upstream of the artery. Net flow per cycle was calculated by integrating the instantaneous flow signal *Q*(*t*) over each 1-second cycle and converted to L/min. Final flow rate at each stage was averaged across 10 consecutive cycles.

### 2.4 Embolic burden quantification

To capture embolic debris, a 100 μm metallic filter (1.5” Buna-N gasket, 150 mesh) was placed downstream near the resistance valve. A fresh filter was installed before each intervention (OA and BA) to isolate debris to the corresponding procedure. After each stage, the filter was photographed under standardized conditions at a fixed distance. A custom Python script (OpenCV-based) segmented embolic particles, excluded background noise by size thresholding, and calculated particle areas (mm^2^) using a calibrated pixel-to-mm conversion. Total embolic area per intervention was then recorded for analysis.

### 2.5 Pulsatility

Arterial pulsatility was assessed under continuous pulsatile flow using duplex ultrasound (DUS) and high-resolution optical recordings. Pulsatility was defined as the relative change between peak systolic and end-diastolic arterial diameters, expressed as a percentage. DUS imaging was performed at baseline, post-OA, and post-OA+BA using a clinical system (Philips CX50, Philips Healthcare, Andover; Figure 2) while maintaining active flow to simulate physiological conditions. In each artery, seven cross-sectional positions were selected for measurement of both inner (ID) and outer diameters (OD), with values averaged to determine mean pulsatility per region. Measurements were taken from representative frames corresponding to peak systole and end diastole using Philips DICOM Viewer R3.0-SP14. Additionally, external arterial motion was recorded using a vertically mounted high-resolution camera (Basler Lens C125-1218-5MP, 12 mm focal length), capturing 10-second video segments per stage. OD measurements were extracted at seven longitudinal locations to compute OD pulsatility, expressed as systolic-to-diastolic diameter ratios.

**Figure 2.**
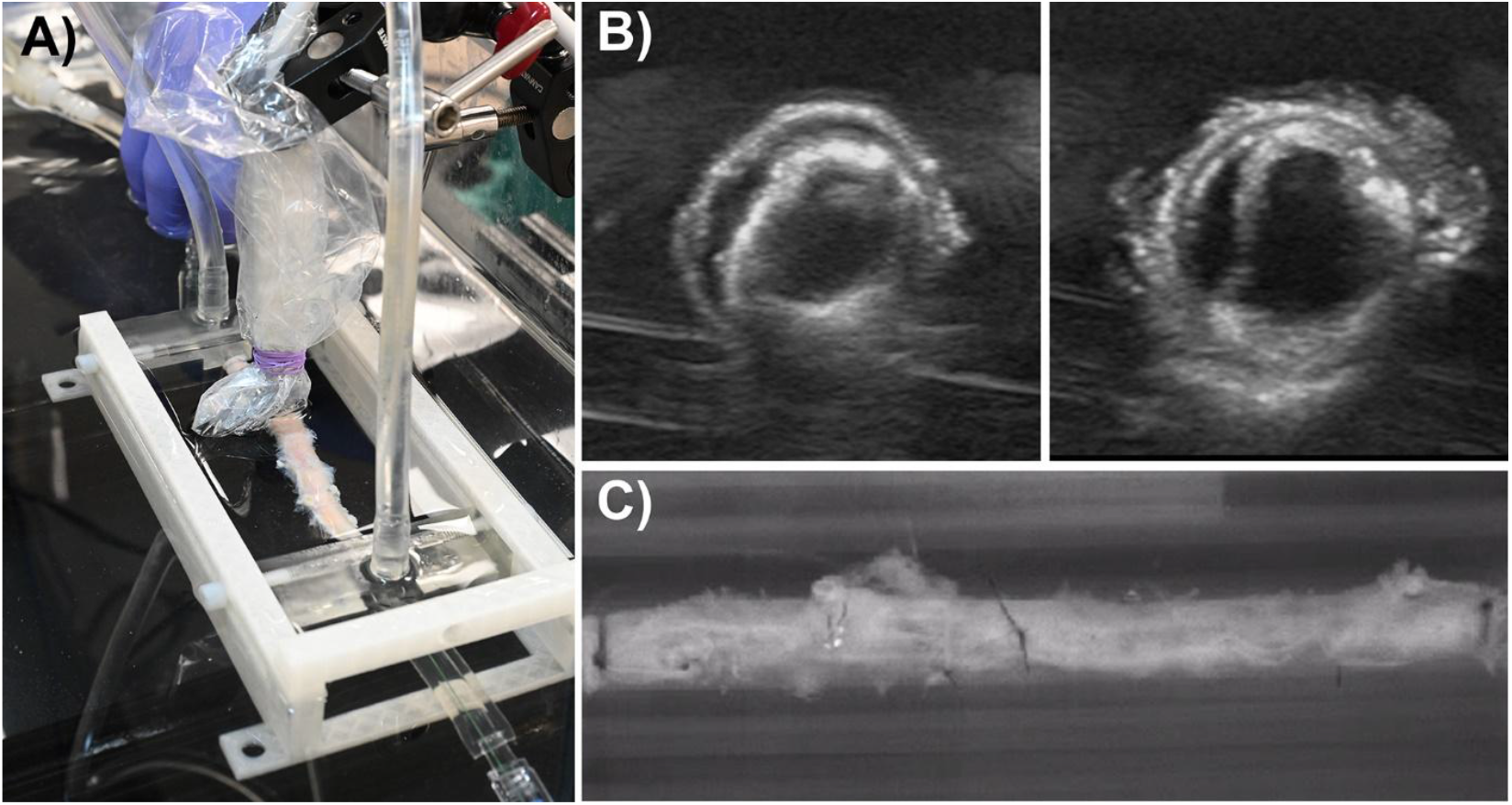
A) Artery mounted in the pulsatile flow circuit with an ultrasound probe positioned for OD pulsatility assessment. B) Representative DUS images after OA showing a post-procedural dissection. C) Top-down view from the camera used to quantify OD pulsatility.

### 2.6 Statistical analysis

To assess the effect of treatment stage (baseline, post-OA, post-OA+BA) on continuous outcomes (e.g., flow lumen volume, translesional mean arterial pressure, net flow rate, and pulsatility), repeated measures ANOVA was performed using SPSS Statistics v29 (IBM Corp., Armonk, NY). Each artery served as its own control, with the within-subjects factor being procedure (three levels). Mauchly’s test assessed sphericity; Greenhouse-Geisser corrections were applied if violated. Multivariate tests (Wilks’ Lambda, Pillai’s Trace) were also reviewed for robustness. Significant overall effects were followed by Bonferroni- or Sidak-corrected pairwise comparisons. Data are reported as mean ± standard deviation, with 95% confidence intervals calculated for marginal means where appropriate. Statistical significance was set at p < 0.05.

## 3. RESULTS

At baseline, the mean arterial stenosis across all specimens was 65% ± 16% (range: 43% to 100%, including one total occlusion). Calcification burden averaged 18% ± 5% of the arterial wall volume, with individual values ranging from 5% to 22%. In 90% of specimens, nodular and plate-like calcifications predominated, while 10% exhibited primarily fragmented or sheet-like morphologies[14].

### 3.1 Volume of the flow lumen

Figure 3 shows a representative μCT image of an artery, illustrating changes in lumen cross-sectional area along its length, with corresponding cross-sections at baseline, post-OA, and post-OA+BA. OA increased lumen size primarily through calcified plaque removal but also induced dissections, which were further exacerbated by BA. Quantitative analysis confirmed a significant and additive increase in flow lumen volume across stages (p < 0.001, partial η^2^ = 0.739). A significant linear trend (p < 0.001) indicated a consistent volume increase with treatment. Post hoc comparisons revealed significant differences between all treatment stages (Figure 6A): OA vs. baseline (Δ = 0.180 mL, p = 0.027, 95% CI [0.021, 0.339]); BA vs. OA (Δ = 0.229 mL, p = 0.024,, 95% CI [0.031, 0.427]); and baseline vs. combined treatment (Δ = 0.409 mL, p < 0.001, 95% CI [0.265, 0.553]).

**Figure 3.**
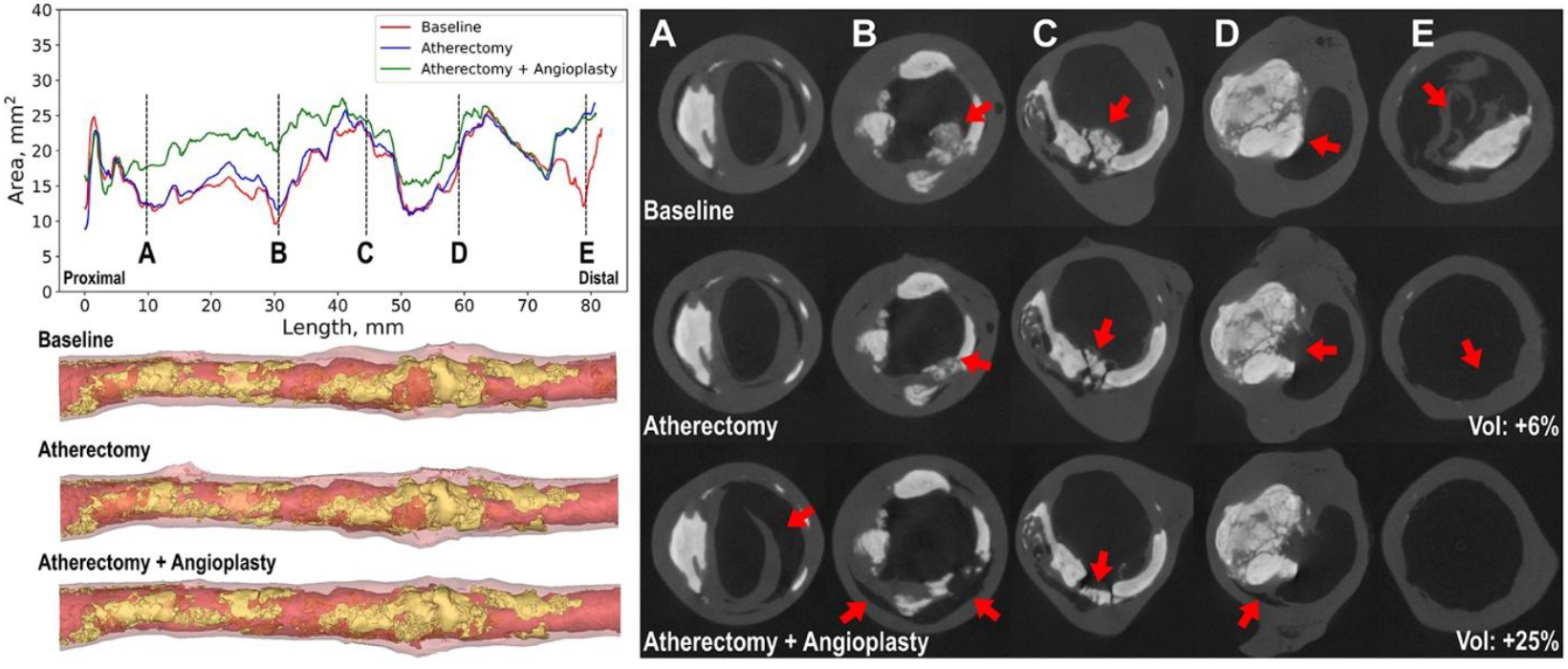
Representative μCT image of an artery, illustrating changes in lumen cross-sectional area along the arterial length after OA and subsequent BA. Red arrows mark treatment-induced changes; “vol” indicates percent increase in flow lumen volume. Panels A–E show sequential cross-sections. OA removed large calcified fragments (later found in the distal filter) and induced dissections, typically at plaque-wall interfaces.

### 3.2 Translesional mean arterial pressure gradient (ΔMAP)

Changes in ΔMAP varied across specimens. In some arteries (Figure 4A), OA produced a substantial reduction in translesional gradient, with further improvement after BA. In others (Figure 4B), ΔMAP showed minimal change post-OA and increased following BA, likely due to distal embolization from large debris. Overall, ΔMAP differed significantly across treatment stages (p = 0.034, partial η^2^ = 0.313, Figure 6B). A clinically meaningful reduction (>10 mmHg) was seen in 40% of specimens, while 50% showed minimal improvement (<10 mmHg), and 10% experienced an increase, likely due to flow-limiting dissection. Despite significant overall ANOVA, post hoc comparisons were not significant (Bonferroni p = 0.143–1.000; Sidak p = 0.116–0.991). Mean ΔMAP decreased from 57.6 mmHg at baseline to 36.7 mmHg after OA and 41.0 mmHg after BA, indicating a general trend toward improved hemodynamics, albeit with considerable inter-specimen variability largely stemming from embolization or dissection.

**Figure 4.**
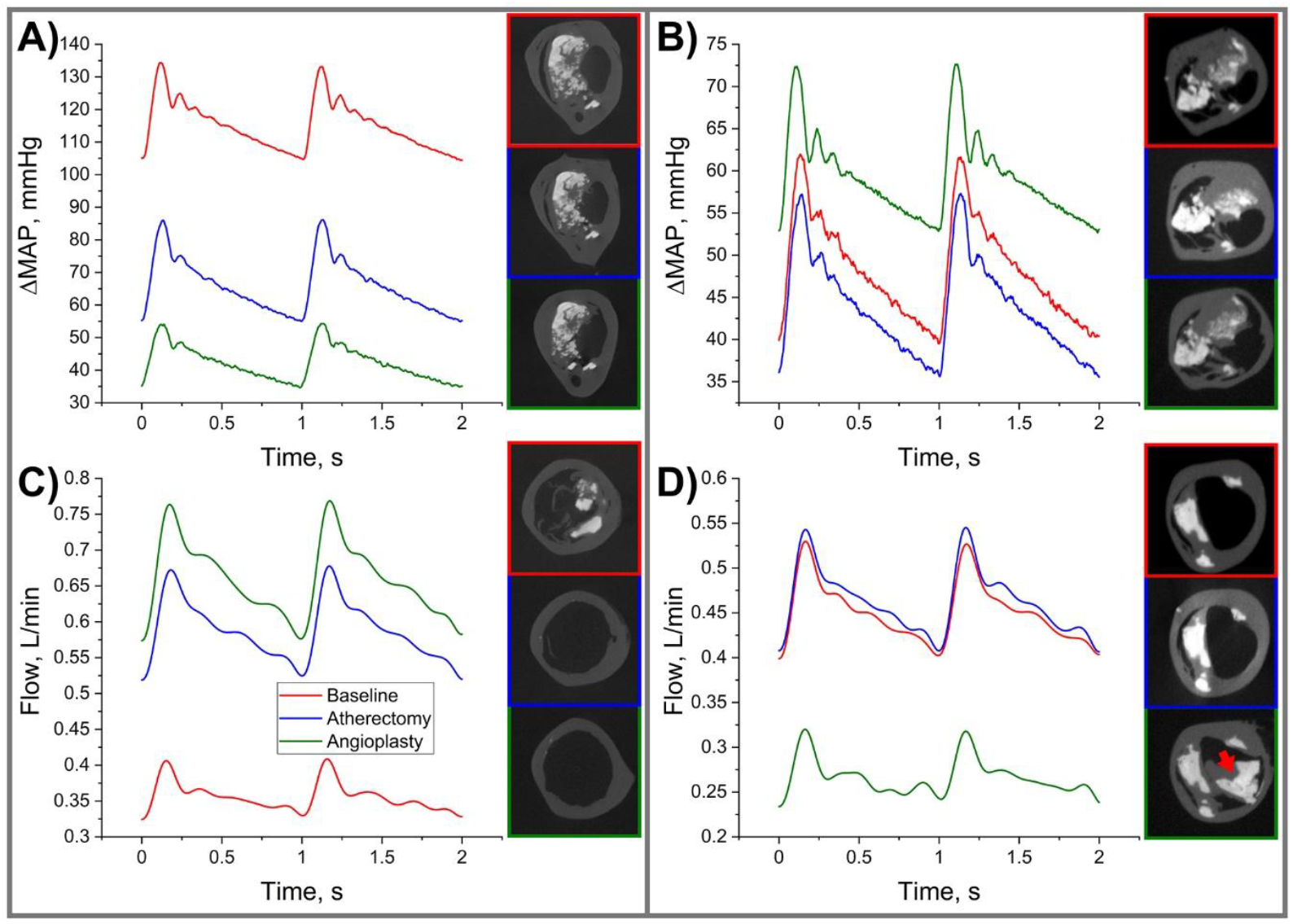
Translesional mean arterial pressure gradient (ΔMAP; A, B) and net flow rate (L/min; C, D) for two representative arteries. In the first case (A, C), both ΔMAP and flow improved after OA and BA. In the second (B, D), ΔMAP showed little change post-OA, and flow worsened after BA due to embolic obstruction from dislodged debris.

### 3.3 Net flow rate

Net flow changes generally paralleled ΔMAP trends, increasing in some specimens (Figure 4C) and decreasing in others (Figure 4D). While the overall effect across treatment stages was not significant (p = 0.126, partial η^2^ = 0.227, Figure 6C), a significant quadratic trend was detected (p = 0.034, partial η^2^ = 0.409), indicating an initial flow increase after OA followed by a plateau post-BA. Clinically meaningful flow increases (≥25%) occurred in 40% of arteries - 30% after OA alone. However, 30% showed no change, and another 30% experienced flow reduction, including one artery with a 41% decrease due to dissection. Mean flow rose from 0.441 L/min at baseline to 0.508 L/min post-OA, remaining stable at 0.502 L/min after BA. No pairwise comparisons were statistically significant (p > 0.12).

### 3.4 Embolic burden

Figure 5 illustrates representative embolic debris captured after OA and BA. OA generated larger fragments (mean area 0.386 ± 1.654 mm^2^; 95% CI: 0.290-0.481 mm^2^; max >30 mm^2^), while BA produced smaller debris (mean 0.143 ± 0.361 mm^2^; 95% CI: 0.096-0.190 mm^2^; max ∼4 mm^2^). Size distributions (right panel, Figure 5) were mapped to diameters of lower limb arteries to indicate potential for occlusion.

**Figure 5.**
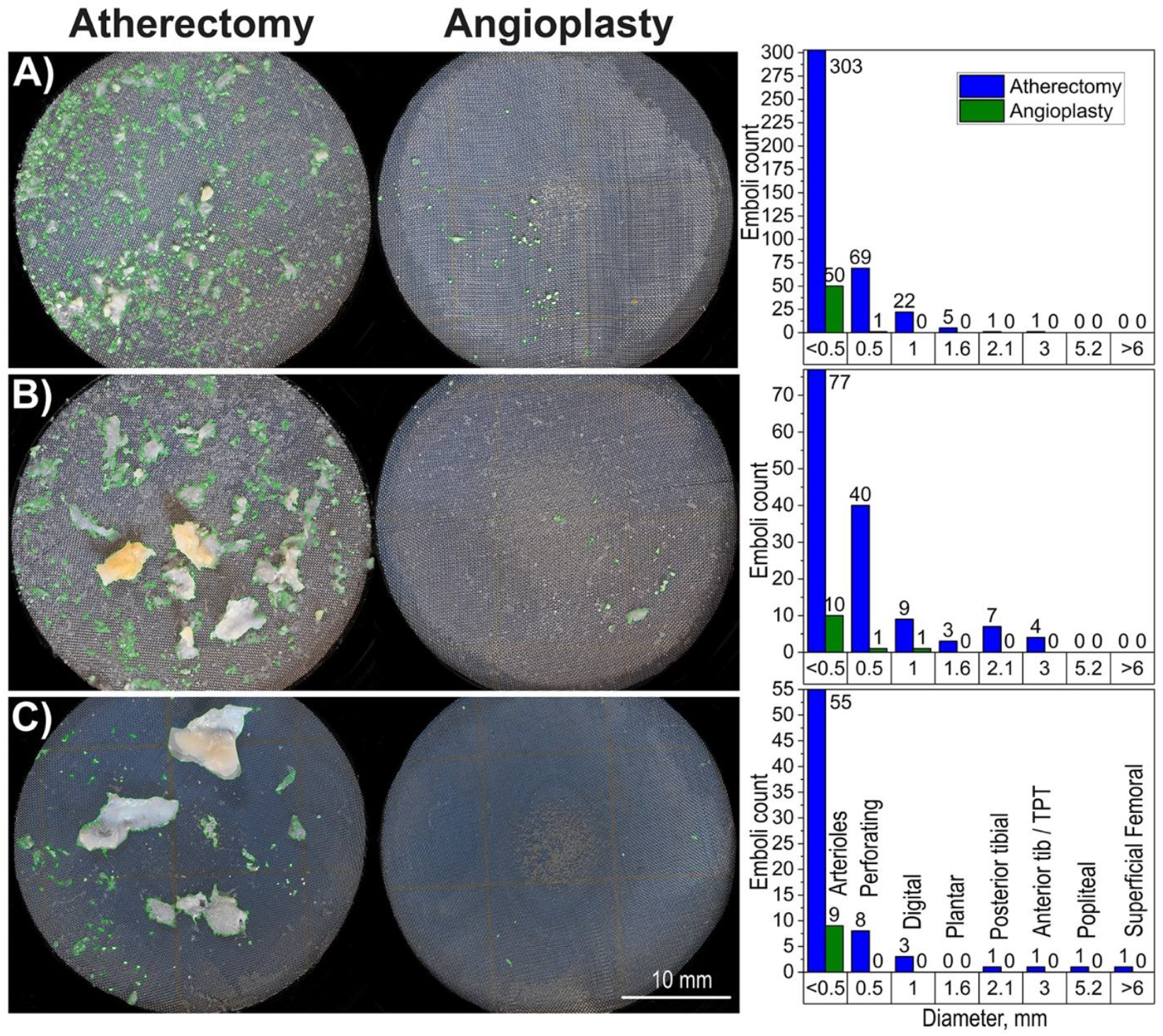
Embolic debris collected after OA and BA in three representative cases (A-C). Green contours outline individual emboli captured on 100 μm filters, used to calculate particle area. Right panels show size distributions based on equivalent diameters (mm), annotated with typical lower limb artery sizes to indicate potential for vascular occlusion.

**Figure 6.**
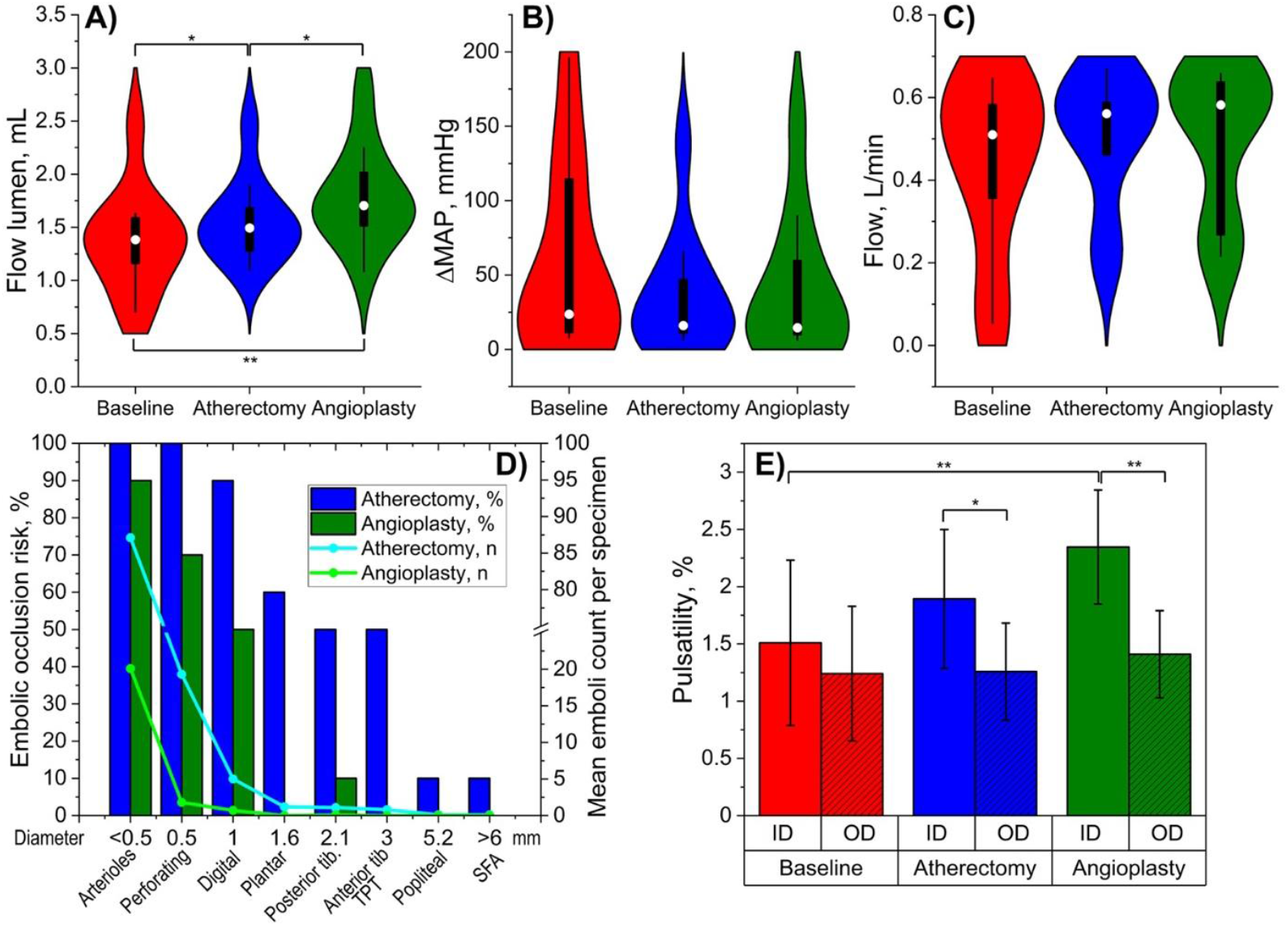
Summary of treatment effects. A) Flow lumen volume (mL), B) ΔMAP (mmHg), and C) net flow rate (L/min) across stages. D) Embolic occlusion risk (%) and mean emboli count per specimen by size; typical lower extremity artery diameters shown for reference. E) Arterial pulsatility (%) by ID and OD via DUS. p < 0.05 (*), p < 0.001 (**).

Figure 6D summarizes embolic risk, with bar graphs showing the occurrence of at least one embolus of a given size and solid lines representing the mean number of emboli per specimen. OA consistently produced emboli capable of occluding arterioles (<0.5 mm, mean 87/specimen), perforator arteries (0.5-1.0 mm, mean 19), and digital arteries (1.0-1.6 mm, present in 90% of cases, mean 5.5). Emboli ≥1.6 mm (plantar and tibial size range) were seen in 50–60% of specimens, with occasional fragments >5.2 mm that could obstruct the popliteal or femoral arteries (10% of cases, mean 0.1). BA generated fewer emboli overall, but still released particles capable of occluding arterioles (mean 20), perforators (mean 1.8), and digital arteries (mean 0.7), likely dislodged from previously loosened plaque.

### 3.5 Arterial pulsatility

Repeated measures ANOVA demonstrated a significant effect of treatment stage on ID pulsatility measured by DUS (p = 0.010, partial η^2^ = 0.400; Figure 6E), with values increasing from 1.510% ± 0.722% (95% CI: 0.994-2.026) at baseline to 1.893% ± 0.662% (95% CI: 1.459-2.327) post-OA, and 2.346% ± 0.498% (95% CI: 1.990-2.702) post-BA. A strong linear trend was observed (p = 0.001), and post hoc analysis confirmed a significant increase from baseline to post-BA (p = 0.004), while other pairwise comparisons were not significant (baseline vs. OA p = 0.738; OA vs. BA p = 0.195).

In contrast, OD pulsatility measured by DUS (Figure 6E) and video imaging did not change significantly across stages (DUS: p = 0.674, camera: p = 0.664). OD values remained relatively stable (DUS: 1.241% ± 0.586% to 1.410% ± 0.380%; camera: 1.523% ± 0.173% to 1.577% ± 0.178%) with no significant pairwise differences. Notably, OD pulsatility was consistently lower than ID pulsatility (Figure 6E); this difference became significant post-OA (p = 0.004) and post-BA (p < 0.001), suggesting external arterial wall motion remained unchanged despite increased luminal compliance.

## 4. DISCUSSION

PAD with severe calcification remains a significant challenge for endovascular revascularization. Atherectomy devices like OA were developed to improve vessel preparation by debulking calcified plaque, increasing luminal gain, and reducing barotrauma during BA. While procedural benefits such as improved compliance and reduced stent use have been reported, clinical studies consistently show higher embolic complications and uncertain long-term outcomes compared to BA or stenting.

Large registries support this concern. In the VSGNE registry, OA had a 4.3% embolization rate - 77% and 64% higher than BA and stenting, respectively[7]. LIBERTY 360 reported 7.8% embolization in diabetic CLI patients treated with OA - four times higher than in non-diabetics[8]. The OASIS trial reported embolic events in 4% of OA cases, though without emergency bypass or unplanned amputations at six months[15]. Our findings align with these: OA generated large embolic fragments (>5 mm) capable of occluding major vessels. Given that embolic protection devices may not filter fragments this large, the clinical implications are considerable. Notably, 68% of embolic events in VSGNE required additional intervention – mostly endovascular (57%), but 11% involved open surgery[7].

Other atherectomy modalities also show high embolic risk. In DEFINITIVE-LE, embolization occurred in 3.8% of directional atherectomy cases[16]. Jetstream, a rotational system with active aspiration, reports 1-8% embolization rates, reduced by embolic filters[17, 18]. A recent Japanese study showed angiographic embolization in 35.5% and true embolic events in 55.4% of Jetstream interventions in calcified FPA lesions[19]. Although reported 1-year limb salvage was 99%, these studies confirm high embolic burden – consistent with our observations.

OA is also thought to restore vessel compliance, enabling low-pressure ballooning and reducing stenting. The COMPLIANCE 360° trial reported a 5.3% bailout stenting rate with OA vs. 77.8% with BA alone[6]. We found a significant increase in ID pulsatility (from 1.5% to 2.3%) after OA and BA, suggesting improved distensibility. However, dissections were common, especially post-BA, highlighting the trade-off between plaque removal and vessel injury. Interestingly, OD-based pulsatility – measured via DUS and external video – did not significantly change, suggesting improved lumen compliance without a corresponding change in wall stiffness. Dissections may also have affected ID measurements via plaque displacement.

Beyond emboli, OA can cause dissections and perforations. In DEFINITIVE-LE, directional devices caused perforations in 5.3% of cases[16]. OA, if misused, can also perforate vessels; coronary data show ∼1% perforation with experienced operators[20], but peripheral rates are less defined. The FDA MAUDE database notes perforation in 2.4% and dissection in 3.4% of 500 Jetstream cases, often requiring reintervention[10]. Embolism (4.4%) was the most frequent complication, underscoring the need for cautious use in heavily calcified or tortuous vessels.

Whether OA improves long-term limb outcomes remains unclear. Some single-arm and registry studies show low amputation rates – for example, OASIS reported no unplanned amputations at 6 months[15], and LIBERTY 360 showed <10% major amputation at 3 years[8]. However, broader analyses suggest no limb salvage benefit. The VQI-Medicare study found a 3.7-fold higher risk of major amputation with atherectomy compared to stenting and a 1.5-fold compared to BA, alongside more major adverse limb events (MALE)[9]. This may reflect more complex lesions treated with atherectomy, but also raises concerns that embolization and dissections may negate its procedural gains. Subgroup analyses offer a more nuanced view: Jamil et al.[21] reported lower amputation and MALE rates for OA+BA vs. BA alone in tibial CLI, albeit with more reinterventions. HCUP data similarly showed reduced in-hospital amputation and mortality with OA vs. BA in CLI patients[22]. These findings emphasize the importance of patient selection – atherectomy may benefit severe calcific CLI but not milder disease.

OA is also associated with high procedural costs. In the U.S., the Diamondback 360 catheter costs $3,795. The average index procedure costs $11,729, with a 2-year PAD-related cost of $29,474-$6,500 or 8,000 more than BA or stenting[23]. Medicare outpatient atherectomy reimbursement rose from $86 million in 2011 to $612 million in 2021[11]. Given this financial burden, its use should be justified by clear clinical benefit, particularly in populations lacking robust evidence.

Our benchtop model provides controlled insight into the hemodynamic and embolic effects of OA, but several limitations apply. The modest sample size (n = 10) limited statistical power and prevented subgroup analysis by plaque morphology or donor risk factors, though findings were consistent for embolic burden and complications. All specimens were cadaveric, lacking vasoreactivity, healing, and thrombosis, which may impact long-term outcomes. The absence of soft tissue may have altered OA crown–wall interaction, possibly underestimating debris generation. While the flow loop mimicked resting FPA hemodynamics, it did not replicate *in vivo* conditions like limb flexion[24] or dynamic pressure changes. Embolic burden was precisely quantified, but downstream perfusion effects were not directly measured. Despite these limitations, the study offers mechanistic evidence for the high embolic burden observed clinically and characterizes OA performance in heavily calcified arteries.

## Data Availability

All data produced in the present study are available upon reasonable request to the authors

## DECLARATIONS

### Ethics approval and consent to participate

This study utilized human donor arterial tissues. Because these donor tissues are de-identified and obtained post-mortem, they do not meet the regulatory definition of human subjects and are therefore not subject to Institutional Review Board (IRB) oversight. Consent for research use of the tissues was obtained from the donors’ next of kin by the accredited organ and tissue procurement organization, Live On Nebraska, as described in the manuscript.

### Clinical trial number

Not applicable.

### Consent for publication

Not applicable.

### Availability of data and materials

The datasets used and/or analyzed during the current study are available from the corresponding author on reasonable request.

### Authors’ contributions

Conception and design: JM, AK

Acquisition of data: RG, MG, BO, PD, JM

Analysis of data: AK, PG

Interpretation of data: AK, JM

Drafting and revising the manuscript: RG, BO, MG, PD, JM, AK

Approval of the submitted version: RG, BO, MG, PD, JM, AK

## Acknowledgements

This work was supported in part by NIH awards HL125736, HL180371, and P20GM152301. The authors thank the Tissue Analysis Core (TAC) of the NIH Center for Cardiovascular Research in Biomechanics (CRiB) and Live On Nebraska for their support, and gratefully acknowledge the tissue donors and their families for making this study possible.

